# Remdesivir induced viral RNA and subgenomic RNA suppression, and evolution of viral variants in SARS-CoV-2 infected patients

**DOI:** 10.1101/2020.11.18.20230599

**Authors:** Florencia A.T. Boshier, Juanita Pang, Justin Penner, Joseph Hughes, Matthew Parker, James Shepherd, Nele Alders, Alasdair Bamford, Louis Grandjean, Stephanie Grunewald, James Hatcher, Timothy Best, Caroline Dalton, Patricia Dyal Bynoe, Claire Frauenfelder, Jutta Köeglmeier, Phoebe Myerson, Sunando Roy, Rachel Williams, The COVID-19 Genomics UK (COG-UK) consortium, Emma C Thomson, Thushan I de Silva, Richard A. Goldstein, Judith Breuer

## Abstract

While changes in SARS-CoV-2 viral load over time have been documented, detailed information on the impact of remdesivir and how it might alter intra-host viral evolution is limited. Sequential viral loads and deep sequencing of SARS-CoV-2 recovered from the upper respiratory tract of hospitalised children revealed that remdesivir treatment suppressed viral RNA levels in one patient but not in a second infected with an identical strain. Evidence of drug resistance to explain this difference was not found. Reduced levels of subgenomic (sg) RNA during treatment of the second patient, suggest an additional effect of remdesivir on viral replication that is independent of viral RNA levels. Haplotype reconstruction uncovered persistent SARS-CoV-2 variant genotypes in four patients. We conclude that these are likely to have arisen from within-host evolution, and not co-transmission, although superinfection cannot be excluded in one case. Sample-to-sample heterogeneity in the abundances of variant genotypes is best explained by the presence of discrete viral populations in the lung with incomplete population sampling in diagnostic swabs. Such compartmentalisation is well described in serious lung infections caused by influenza and *Mycobacterium tuberculosis* and has been associated with poor drug penetration, suboptimal treatment and drug resistance. Our data provide evidence that remdesivir is able to suppress SARS-CoV-2 replication *in vivo* but that its efficacy may be compromised by factors reducing penetration into the lung. Based on data from influenza and *Mycobacterium tuberculosis* lung infections we conclude that early use of remdesivir combined with other agents should now be evaluated.

**Summary Sentence:** Deep sequencing of longitudinal samples from SARS-CoV-2 infected paediatric patients identifies evidence of remdesivir-associated inhibition of viral replication *in vivo* and uncovers evidence of within host evolution of distinct viral genotypes.

## Introduction

Severe acute respiratory syndrome coronavirus 2 (SARS-CoV-2), which causes Coronavirus Disease 2019 (COVID-19), was first identified in Wuhan, China in December 2019. On 11th March 2020, the WHO declared COVID-19 a global pandemic *(1)*. Since then, an estimated 50 million people have been infected of whom up to 2.5% have died *(2)*. A number of studies have assessed nasal and oropharyngeal viral load data from longitudinally sampled SARS-CoV-2 infected patients. Their findings reveal wide variations in viral load at presentation *(3– 6)*. However, milder disease and clinical recovery are associated with lower and declining viral load respectively, pointing to its potential use as a biomarker for antiviral drug response. Remdesivir, an RNA-dependent-RNA polymerase (RdRp) inhibitor, has been shown in one large randomised clinical trial (RCT) to be effective against SARS-CoV-2, although another large study, showed no clinical benefit and smaller studies have shown limited or no impact on clinical recovery *(3,7–11)*. Where clinical trial data are lacking or contradictory we have previously used deep pathogen sequencing, mutational analysis and evolutionary modelling to gain insight into the impact of repurposed drugs, including RdRp inhibitors similar to remdesivir, on serious respiratory RNA viral infections in hospitalised patients *(12,13)*. These studies have revealed drug-related mutational signatures, evidence of viral compartmentalisation in the lung and previously unrecognised synergy between combination therapies associated with changes in viral loads and improved clinical outcomes. Here we report the application of similar methods in a personalised-medicine-approach to investigating the impact of remdesivir on SARS-CoV-2 within an individual and to evaluate potential biomarkers that can be used to monitor clinical efficacy. The data provide further insights into fundamental questions of SARS-CoV-2 evolution and co-infection.

## Results

### Overview of Patients

We analysed nine hospitalised SARS-CoV-2 positive paediatric cases with a mean age at time of infection of 4.7 years old (range 0-14 years old), who were repeatedly sampled during the course of their SARS-CoV-2 infection. A summary of their clinical features is shown in Table 1. Five patients had pre-existing co-morbidities associated with primary or secondary immunodeficiency (Table 1). Of the nine patients, four (A, D, G and H), of whom only H was known to be immunocompromised, were admitted to the Paediatric Intensive Care Unit (PICU) (supplementary figure 1). The remaining patients were cared for in appropriate source isolation and were not co-located during their SARS-CoV-2 infection (supplementary figure 1). Patients, A, D, and G received 8 - 10 days of remdesivir through the compassionate access programme. Patient A and G received 200 mg loading dose followed by 100 mg daily and Patient D received 5mg/kg (10 mg) loading dose and 1.25 mg/kg (2.5 mg) once daily (Table 1).

**Table 1:** Overview of patient clinical, treatment, sampling and viral lineage data for all 9 patients.

| Patient | Gender | Age Group (Years) | Ethnicity | Weight (kg) | Diagnosis | Immune Status | Health Care Associated | Days from first recorded symptom to first positive | Remdesivir (Y/N) [days] | Days in ICU | No. of Sequenced Samples | Lineage | Previous Studies |
| --- | --- | --- | --- | --- | --- | --- | --- | --- | --- | --- | --- | --- | --- |
| A | Female | 6-10 | Other | 39 | bronchiectasis with prior lobectomy | Immunocompetent |  | 3 | Y [8] | 12 | 9 | B.1.1 | (76–78) |
| B | Female | 6-10 | Asian | 25.7 | severe chronic illness including kidney disease | Immunocompromised |  | Unknown | N | 0 | 3 | B.1.1 | (76–78) |
| C | Male | 6-10 | Black African | 52.2 | lymphoblastic leukaemia, on chemotherapy | Immunocompromised |  | 0 | N | 0 | 5 | B.2.1 | (76–78) |
| D | Male | 0-1 | White | 1.98 | ex 32 week prematurity, required inotropes and ventilation | Immunocompetent |  | 1 | Y [10] | 22 | 7 | B.1.1 | (76–79) |
| E | Male | 2-5 | Other | 12.4 | Neuroblastoma on chemotherapy | Immunocompromised | Suspected | 1 | N | 0 | 5 | B.1.p16 | (76–78) |
| F | Male | 0-1 | Unspecified | 7.4 | extreme prematurity, short gut, chronic lung disease | Likely immunocompromised | Suspected | 0 | N | 0 | 2 | B.2.1 | (76–78) |
| G | Male | 11-15 | Unspecified | 120 | obese | Immunocompetent |  | 10 | Y [10] | 10 | 10 | B.1.1.7 | (76–78) |
| H | Male | 0-1 | Black African | 10.3 | Hepatoblastoma | Immunocompromised |  | Unknown | N | 34 | 13 | B.1.p11 | (76–78) |
| I | Female | 0-1 | Unspecified | 9.85 | PNET, on chemotherapy | Immunocomrpromised | Suspected | Asymptomatic | N | 0 | 2 | B.1.p16 | (76–78) |

### Viral load trajectories by cycle threshold and clinical markers of infection

Figure 1 shows the viral polymerase chain reaction (PCR) cycle threshold (ct) values for all nine patients for forty days following their first positive nasopharyngeal aspirate (NPA) available to us. Viral RNA was measured in NPAs for all patients and/or upper airway secretions for those who were intubated (patients A, D, G, and H). Patients A also had a Bronchoalveolar lavage. In agreement with earlier studies *(14)*, the ct values showed considerable day-to-day variation of between 0.16 and 14.4 cycle numbers (median 5.5 cycle numbers). Viral RNA continued to be detectable for 7 to over 50 days (median 16 days) following the first positive sample (supplementary figure 2). Of the three patients who received remdesivir, only patient D had total suppression of viral RNA during treatment followed by rebound of virus after treatment cessation (figure 1). The four ICU patients were clinically most unwell, requiring assisted ventilation. All three remdesivir-treated patients showed clinical improvement after starting the drug, associated with falls in temperature (all) and inflammatory markers (A and G) (supplementary figure 3). All three were weaned from conventional ventilation before the treatment course was completed with decreases in oxygen requirements. In patient D, a significant reversal of respiratory deterioration was noted once remdesivir was started; inhaled nitric oxide was stopped within 96 hours coincident with weaning from high frequency oscillatory to conventional ventilation. Patient D, who alone required inotropic support, achieved haemodynamic stability off inotropes within 5 days of starting remdesivir.

**Figure 1.**
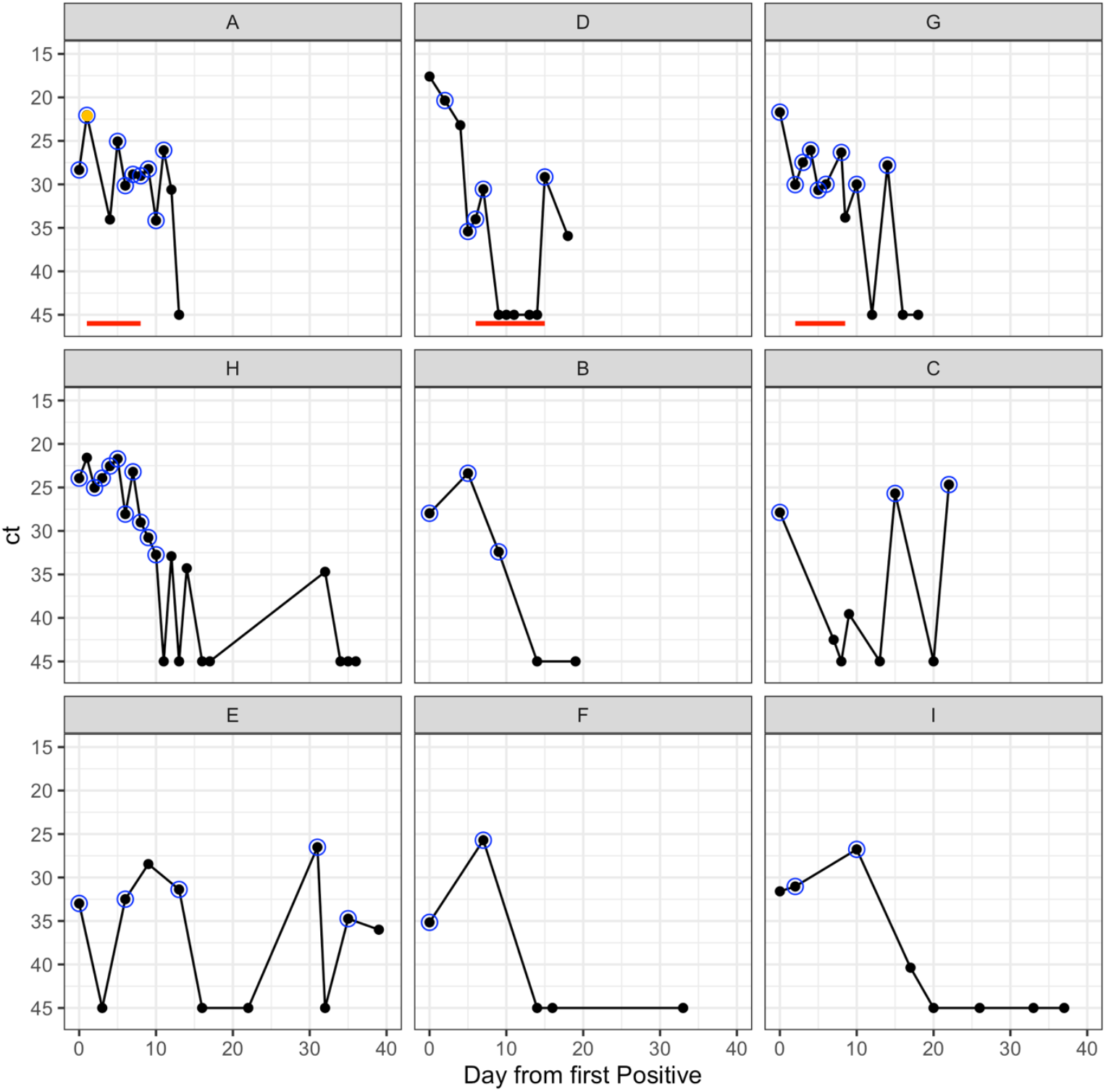
Ct trajectories of 9 patients from first day up to 40 days post first positive. One panel per patient, red line indicates remdesivir received, black dot is sample taken, blue circle indicates sample successfully sequenced. Orange dot indicates Bronchoalveolar lavage sample.

### Inter and intra-host phylodynamics of SARS-CoV-2

To investigate the possibility of remdesivir resistance in patients A and G, neither of whose viral RNA was suppressed by remdesivir treatment (figure 1), and in patient D whose virus rebounded following cessation of treatment, we deep sequenced all samples as well as those from untreated patients for comparison. The sequencing metrics for the samples are summarised in Supplementary Table 1. Relative to their first available sample, there were nine polymorphisms identified in viruses from patients A, H and I of which five were non-synonymous with four in the ORF1ab (nsps 1,3,4,5) and one in the Spike protein, S2 subdomain (supplementary table 2). This latter mutation P812L in patient I is located in a T-cell epitope *(15)*. None of the identified SNPs were sites identified as common homoplasies or those known to be susceptible to Illumina sequencing error, none have been associated with remdesivir resistance and none other than P812L were in known or predicted immune epitopes *(16–22)*. Both consensus level mutations in patient A, who received remdesivir were non-synonymous. One was located in the nsp4 (responsible for formation of double membrane vesicle) and the other in nsp5 (a chymotrypsin-like protease responsible for polypeptides cleaving at 11 sites downstream to release nsp4-nsp16 and inhibition of interferon signalling) of ORF1ab *(18,23,24)*. Nsp5 has been shown to mediate the maturation of nsps downstream which is an essential part of the viral lifecycle *(23)*; this has made it a desirable target for antiviral drug production *(25,26)*. However, the nsp5 mutation P241L observed in patient A was not found to be in an active site. Virus from patient D, which rebounded following cessation of remdesivir treatment, was genetically identical to pre-treatment virus.

Patients H and I who did not receive remdesivir, also had consensus level sequence changes (supplementary table 2). Only one of four mutations for patient H was non-synonymous and this was in nsp3 (a papain-like protease) in ORF1ab. Two of three mutations in patient I were non-synonymous, one in nsp1 (responsible for cellular mRNA degradation and inhibition of interferon signalling) in ORF1ab and the other upstream of the furin cleavage site and the S2’ cleavage site in the spike protein *(27,28)*. This Spike mutation although not predicted to alter structure, does lie within a known CD4 epitope although it is not known whether it would abrogate T cell binding (supplementary figure 4) *(15)*. Patient E was negative for SARS-CoV-2 on consecutive samples obtained on days 16 and 22. Virus detected again at days 31 and 35 was identical to all other sequences from this patient. We observed changes in the consensus sequences between different samples from patients A, H and I. Sequences A at time point 6, 7, and 8 were identical to those of samples sequenced from patient D as depicted in Figure 2 A. No evidence of laboratory contamination to explain the identity between A and D sequences was found, with samples from these patients being sequenced in separate batches on separate days.

**Figure 2:**
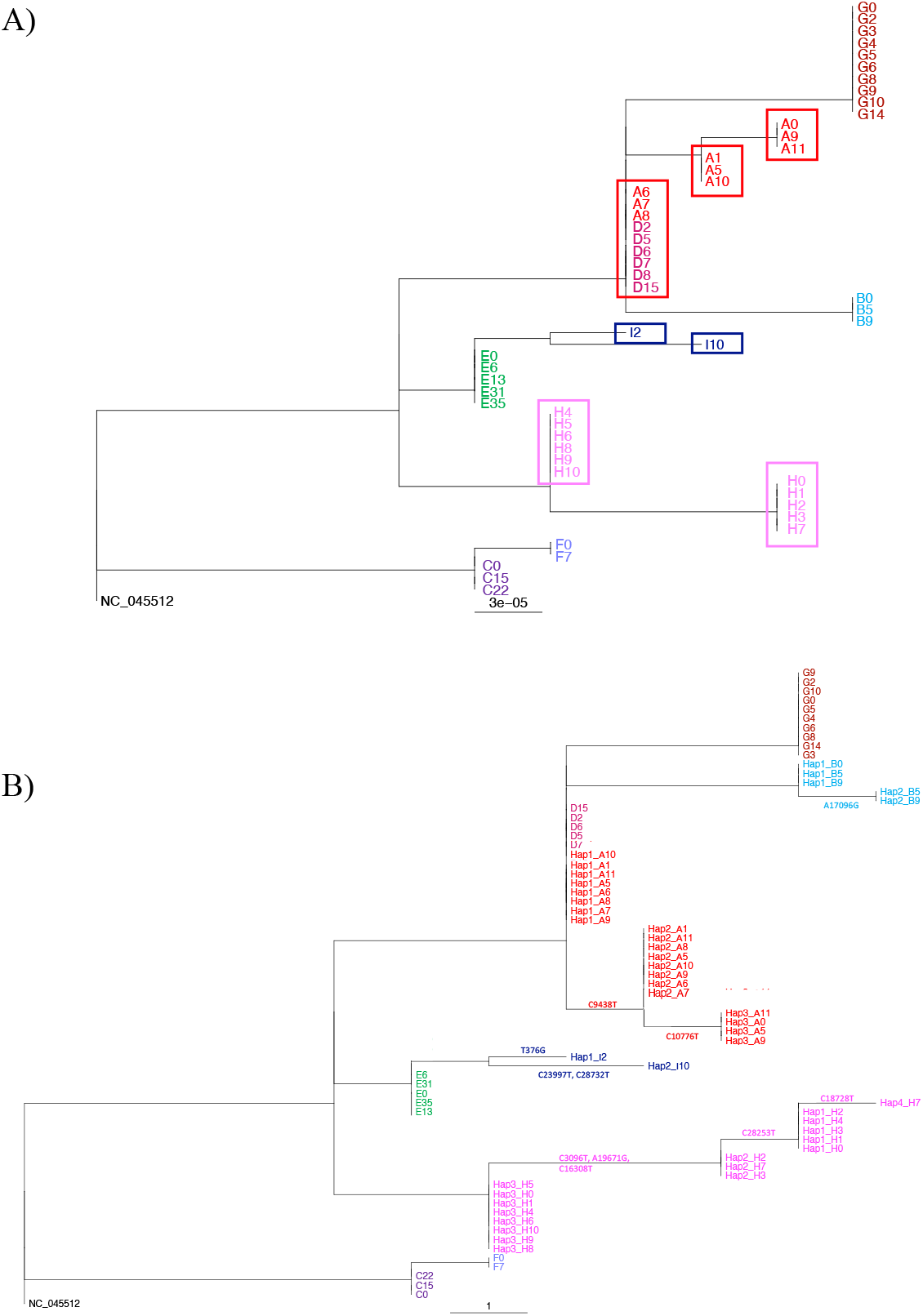
RAxML phylogenetic trees rooted at NC_045512. **A)** Tree using consensus level sequences for Patients A-I. Boxes highlight distinct, identical sequences excluding gaps, found in patients A, H and I over time. Samples are labelled as [Patient][Time]. **B)** Tree using haplotype sequences for Patients A, B, H, and I and consensus levels sequences for Patients D-G for which no haplotypes are identified. Haplotypes defining mutations are shown along the corresponding branches. Samples are labelled as Hap [number]_[Patient][Time].

### No mutagenic signature identified for remdesivir

Ribavirin and favipiravir, which like remdesivir are nucleoside analogues, have been shown to inhibit RdRps at higher drug concentrations while at lower concentrations they appear to drive the accumulation of mutations, in particular C to T and A to G transversions *(13,29)*. The mutagenic effect is associated with loss of infectivity and with an effective clinical response *(29)*. In vitro studies have not shown lethal mutagenesis to be a feature of remdesivir *(30,31)*. However, to exclude non-lethal mutagenesis as a possible explanation for continuing high viral RNAs despite remdesivir treatment, we compared the mutational burden and patterns of transitions and transversions in treated and untreated patients. In accordance with current understanding of its mode of action, we found neither an increased mutational burden in remdesivir-treated patients nor any evidence of associated mutational signature (supplementary figure 5 and supplementary figure 6). We found no evidence that the proportion of transitions and transversions distribution of mutation was dependent on the treatment (Fisher t-test, p=0.13).

### Measurement of subgenomic RNA

To determine whether, despite stable viral ct values, remdesivir treatment may have inhibited viral replication we analysed subgenomic RNA (sgRNA) detected by the genome sequencing using the program Periscope *(32)*. sgRNA derives from negative stranded RNA intermediates generated during SARS-CoV-2 replication *(33,34)*. SARS-CoV-2 elaborates ten canonical transcripts, although ORF 10 sgRNA has never been detected *(32,35–37)*. To optimise recovery, we excluded genomes with <90% coverage and <100 mean read depth (MRD). Stacked bar-plots of the frequency of sgRNA reads per 100,000 mapped reads (sgRPHT) for each gene and corresponding ct values for each patient are compared in figure 3A. Unlike previous reports, no correlation was found between sgRPHT and ct-values (supplementary figure 7A) *(35)*. We have excluded patient D from the comparative analysis below as their virus was suppressed below the limit of detection during remdesivir treatment, which does not permit us to measure sgRPHT by the current technique. sgRNA levels in samples taken during remdesivir treatment for patients A and G were lower than in samples off treatment (Mann-Whitney-Wilcoxon Test p = 0.05) (figure 3B). Samples 6,7 and 8 in patient A had a mean viral ct values of 28.7 (range 28.24 – 29.02) with barely detectable sgRNA despite high MRDs (3402-6297). sgRNA was detected in all other samples with ct values <35 bar two. A similar tendency towards significance of remdesivir treatment on levels of sgRPHT was shown across all patients over all time points (Mann-Whitney-Wilcoxon Test p= 0.059) (supplementary figure 7 B).

**Figure 3:**
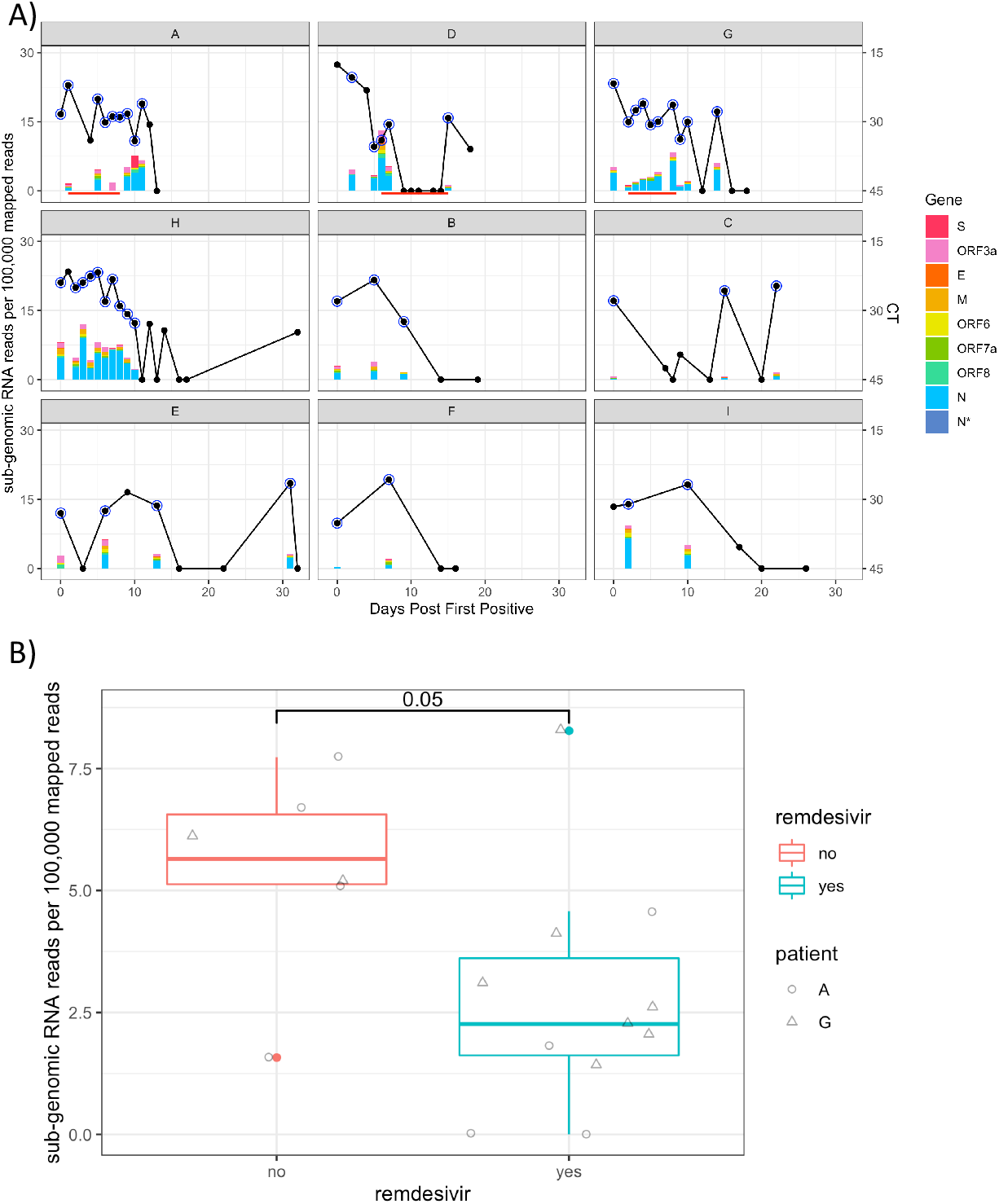
Evaluation of effect of remdesivir on levels of sgRPHT. A) Comparison of ct values and sgRPHT over time by Patient. Stacked bars represent sgRPHT values coloured by gene. Black line represent ct values, with blue circles annotating successfully sequenced samples. Y-axis is days post first positive. B) Box-plot of sgRPHT on and off remdesivir for patients A and G. Samples from each individual are identified by their shape. Treated samples have low sgRPHT than untreated samples taken in the same time-window post first positive (Mann-Whitney-Wilcoxon test, p = 0.05). Patient D was excluded from this comparison as no sequences were available during remdesivir treatment as viral load was below the limit of detection.

### Evidence of mixed infection

We observed changes in the consensus sequences at different timepoints in patients A, H and I as depicted in figure 2A. To examine this and the identical sequences from patients D and A further, we analysed minor variant alleles (MVAs) as outlined in the methods. A complete list of identified polymorphisms for each sample, and corresponding frequency and read support, can be found in supplementary table 3. Patients A, B, H, and I had well supported MVAs which varied in frequency over time (supplementary figure 8). MVAs in other patients occurred for the most part on a single occasion or at levels <20% with poor read support. To resolve possible mixed infections within each sample, we used the haplotype reconstruction method HaROLD *(38)*. HaROLD identified three haplotypes for patient A, four for patient H, two for patient I, two in patient B with one in all other patients. All identified haplotypes are labelled as Hap_[number]_[Patient][Time]. Haplotypes clustered phylogenetically by patient other than for Hap1_A, found at the root of the clade from patient A, which was identical to viral sequences from patient D (figure 2B). Haplotype-defining sites are indicated along branches, the corresponding read support for each of these is identified in supplementary table 3. The abundance of each haplotype over time is shown for patients A, B, H, and I (figure 4 A). We observed no obvious pattern of haplotype change in any of the patients, with sample-to-sample variation in viral loads and haplotype abundance occurring particularly in patients A and H.

Since independent SARS-CoV-2 lineages that differ by one or two SNPs are frequently seen in the UK, we first determined whether the haplotypes within patients A, H, I, and B were likely to represent co-infections with different viruses *(39)*. Using Local Lineage and Monophyly Assessment (LLAMA) *(40)*, we identified the lineage in the global alignment on COVID-19 Genomics UK (***COG***-***UK***) *(41)* consortium nearest to each haplotype. The local trees identified by LLAMA are shown in supplementary figure 9, with a single local tree representing each of the observed lineages. Comparison of haplotypes in patients A, H, and I with global sequences confirmed that haplotypes Hap1_A, Hap2_A, Hap3_H and Hap1_I were circulating as independent lineages in the UK and globally as were the single genotypes from patients C, E, and F (Supplementary Figure 9). However, haplotypes Hap3_A, Hap1/2_B, Hap1/2_H and Hap2_I as well as the virus from patient G were not represented among 61740 global sequences available on GISAID *(42)*. This may reflect incomplete population sampling or the recent emergence of new lineages. Alternatively, it is possible that some haplotypes e.g. Hap3_A, Hap1/2_H and Hap2_I are not currently freely circulating in the population and possibly that the mutations they carry are deleterious as has been postulated for influenza*(43)*. The close clustering between each of the non-circulating viruses with other haplotypes in their cognate hosts supports the possibility of within-host evolution (supplementary figure 9).

**Figure 4:**
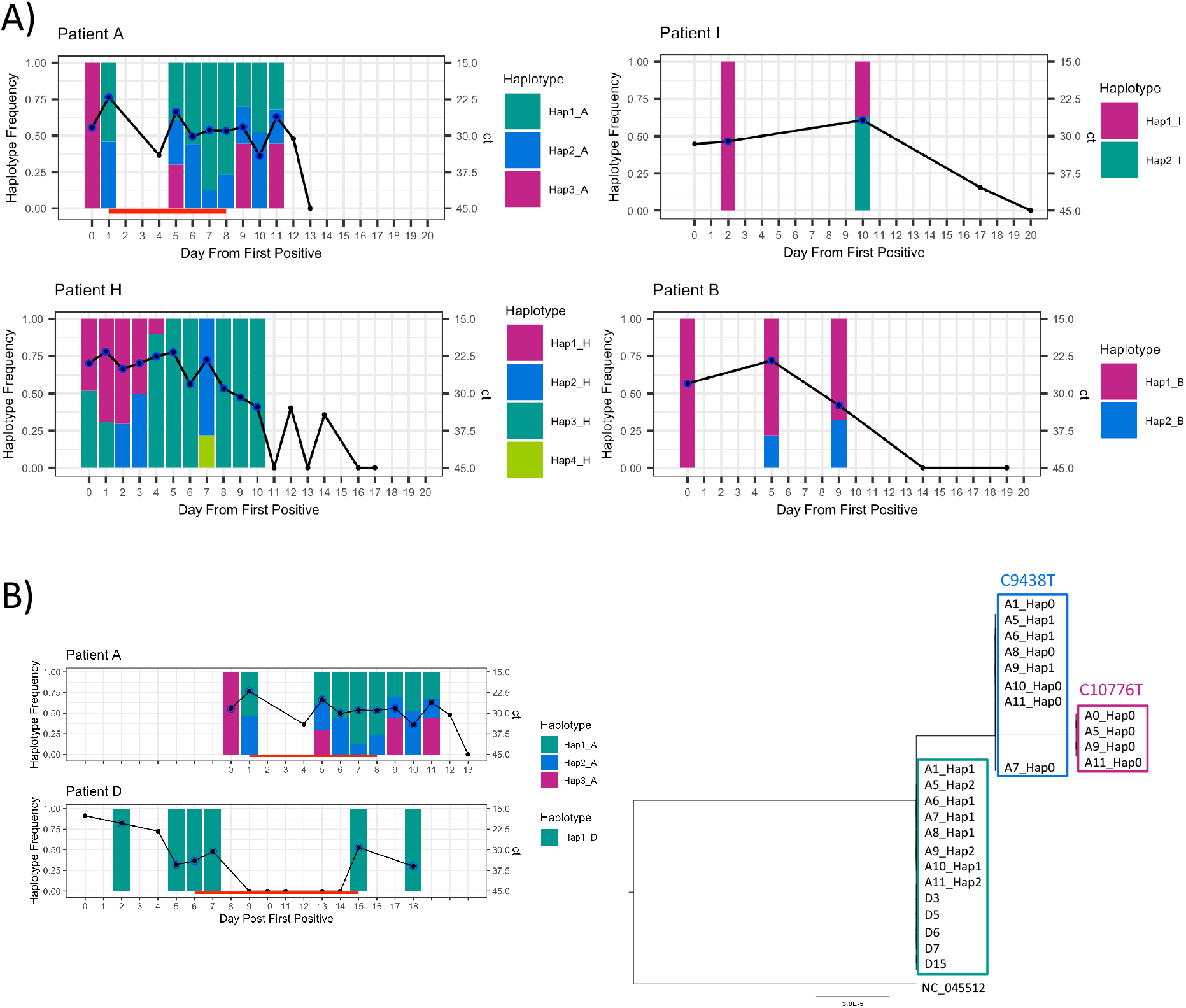
Frequency of identified haplotype over time for individual patients. **A)** Haplotype frequency over time for patients A, B, H and I. **B)** *right:* Phylogenetic tree of haplotypes from patients A and D, nucleotide mutation shown above each cluster *left:* Haplotype frequency over time for patients A and D. Black line is Ct value, red line indicates remdesivir received, black dot is sample taken, blue circle indicates sample successfully sequenced. Bars indicate frequency of identified haplotypes.

An alternative explanation, given that patients A, B, H, and I apparently had mixed populations on first testing or soon after, is that these patients were co-infected with multiple viruses as has previously been suggested *(44)*. Analysis of a separate dataset of 318 sequences collected in Glasgow between 28/02/2020 and 29/04/2020 found no relationship between the number of shared MVAs and the consensus sequence pairwise distance, suggesting that shared diversity is not commonly due to co-transmission in contrast to earlier work (supplementary figure 10) *(44)* As proof of concept we reconstructed haplotypes for four of the sequences, each of which had optimal MVA distribution (as detailed in the methods) for HaROLD analysis. We found between two and three closely related haplotypes for each patient clustered by patient as described for samples A, B, H, and I (supplementary figure 10). As was the case for A, H and I, only one haplotype per patient was represented among freely circulating viruses in global sequence alignment provided by COG-UK, reinforcing the likelihood that the variant genotypes had arisen within each host.

A possible exception to this conclusion was patient A. It is conceivable that haplotype Hap1_A, which is identical to the viral sequence from patient D, could have resulted from superinfection. Patients A and D were co-located in the Intensive Care Unit (ICU) (supplementary figure 1). Haplotype Hap1_A was not identified in the sample taken from patient A before their transfer to the ICU but was detected approximately 28 hours later in a sample taken two hours before starting remdesivir (Figure 4 B). Were superinfection the explanation, the appearance of Hap2_A could be explained either by recombination between Hap3_A and Hap1_D following transfer of patient A to the ICU or by within-host mutation of Hap 3_A. However, no other healthcare associated transmissions were reported in the ICU and local epidemiological investigation made it unlikely. Moreover, in view of the rapid appearance of Hap1_A following transfer to ICU and the variation in haplotype frequencies over time, it is perhaps more probable that Hap1_A and 2_A were present in the patient’s first sample but not detected. This would support within host evolution of these three closely related strains, for one of which, Hap3_A, there is no evidence of circulation in the population.

## Discussion

Our comparative analysis of longitudinal samples from remdesivir-treated and untreated patients infected with SARs-CoV-2 identifies evidence of remdesivir-associated suppression of viral RNA and sgRNA *in vivo* and uncovers the presence of mixed viral haplotypes likely to have evolved within each host early in infection, persisting thereafter, in some cases possibly within discrete tissue compartments in the lung.

Suppression of viral replication is a recognised goal of SARS-CoV-2 treatment, since better clinical outcomes have been associated with lower viral loads *(14,45)*. While several drugs with *in vitro* activity against SARS-CoV-2 have been identified, only remdesivir has shown any clinical promise*(10)*. However, reports of its efficacy are mixed *(3,7–9,11)* and only one other report of it having an effect on viral replication *in vivo* has been described *(46)*. We show evidence of clinical improvement in all three patients whilst on remdesivir. In patient D clinical improvement was associated with a fall in viral RNA levels within two days of starting remdesivir. Viral RNA remained undetectable until treatment was stopped, when it again rose. In patient A no change in viral RNA levels occurred during remdesivir treatment, however, SARS-CoV-2 sgRNA levels appear to have been reduced, increasing again following cessation of treatment (figure 4). sgRNA levels have been shown to have a weak association with viral replication in *in vitro* culture *(47)*, however the extent to which sgRNA levels, mainly of the E gene which are most often measured, reflect viable virus better than viral RNA remains controversial *(35)*. Overall, for samples obtained during remdesivir therapy we observe lower sgRNA levels even in the presence of high viral RNA (figure 4). These samples have good coverage and MRDs (supplementary table 1), thus excluding RNA degradation *(32)*. Our results mirror findings from the macaque model wherein animals are treated with remdesivir 12 hours following infection with SARS-CoV-2. In this model the viability in *in vitro* culture of virus from upper respiratory samples is decreased and clinical scores are improved, despite no change in viral RNA levels *(48)*. The possibility that sgRNA, may, together with viral load, provide a biomarker of response to remdesivir should now be explored.

We have previously used deep sequencing and evolutionary modelling of influenza and norovirus genomes to gain insight into the *in vivo* action and efficacy of ribavirin and favipiravir, drugs with similar modes of action to remdesivir *(13,29)*. Using a similar approach, we show considerable variation in the consensus SARS-CoV-2 sequences obtained from four of the nine patients, which appears unrelated to remdesivir treatment. The heterogeneity is explained by the presence of between 2-4 stable viral haplotypes that vary in abundance (figures 4A). From this we see that, while remdesivir suppressed viral replication in patient D, the same dose had little impact on the viral RNA from the identical strain, Hap1_A, in patient A (figure 4B). Remdesivir resistance was not found to account for the difference in response. Instead, the variable abundance of multiple distinct haplotypes present in different samples from patients A, B, H, and I, including in samples obtained from deep within the lung, is consistent with the presence of tissue compartmentalisation, wherein a pathogen replicates in physically separated niches within the lung potentially accumulating mutations that allow different populations to be distinguished. Diagnostic specimens that incompletely sample these poorly mixed virus populations, result in mixed haplotypes that vary in frequency from sample-to-sample, as demonstrated in our patient data. The findings are supported by the independent finding that SARS-CoV-2 viral loads vary in different areas of lung sampled post mortem *(49)*.

Tissue compartmentalisation is well described for other inflammatory lung infections including *Mycobacterium tuberculosis* (M.tb) and influenza and has been associated with uneven drug penetration, leading to poor resolution of infection and predisposing to drug resistance *(29,50,51)*. Modelling of drug levels within lung tissue from SARS-CoV-2 infected patients suggests that remdesivir penetration into lung tissue is poor *(52)*. In patient A, the persistence of different viral populations at variable frequencies including in bronchoalveolar lavage material supports the presence of viral compartmentalisation. Other patients may also have had tissue compartmentalisation but, in the absence of distinct viral populations, this could not be detected. Another possibility is that the effective remdesivir dose was too low, for example in patient G who was obese (table 1) or that duration of treatment too short, as in patients A and D in whom viral sgRNA and RNA respectively rebounded at the end of treatment (figures 1 and 3A). Clearance of remdesivir metabolites is affected by renal function *(53)*. Neonates, such as patient D, are known to have relatively reduced renal clearance of drugs; accumulation of metabolites may therefore have contributed to viral RNA suppression in patient D *(54)*. Poor drug penetration into infected lung tissue has clearly been demonstrated to underlie suboptimal responses to treatment of M.tb infections *(50,51)* and tissue compartmentalisation is recognised to contribute to the emergence of drug resistance in both Mtb and Influenza *(12,50)*. For both M.tb and Influenza, combination therapies have been shown to achieve better pathogen clearance from lung and clinical outcomes than single agents *(50,55)*.

High fidelity deep sequencing of multiple samples from the same patient together with computational methods that discard poorly supported MVAs minimised the contribution of artefact for the interpretation of within-host diversity in our sample set *(17,38)*. Many of the persistent MVAs were present in early samples. As seen in other RNA viruses MVAs in SARS-CoV-2 may be more common in patients with primary or secondary immunodeficiency *(13,29)*; we did not see the accumulation of mutations over time that characterises, for example, norovirus infections in the immunocompromised (supplementary figures 5 and 6). Haplotypes occurring within an individual were phylogenetically sister taxa, with generally only one haplotype per patient identical to freely circulating lineages (supplementary figure 9). Many circulating SARS-CoV-2 lineages differ by similar numbers (one to two) of SNPs as the haplotypes found within our patients, making it possible that the latter were in fact independent strains that were too rare to have been previously sampled. Such strains would have to have been acquired through co–transmission or superinfection. Although this is possible for patient A, on balance the data do not support this. Another possibility is that mixed haplotypes represent co-transmission of different strains *(56)*. To explore the likelihood of this scenario, we used a large, independent and well curated data set of 318 geographically co-located SARS-CoV-2 sequences. We show that sequences that were phylogenetically closer did not share more MVAs (supplementary figure 10), suggesting the co-transmission of infections is unlikely. The result was not changed by excluding strains sharing single MVAs which are the most likely to be artefact. Analysis of four randomly chosen samples with multiple well supported MVAs at levels of 15-85% (optimal for haplotype reconstruction), generated identical findings to those of our patients, namely sister taxa within hosts with only one viral sequence per patient found among freely circulating lineages. We conclude that early within-host evolution most logically explains the diversity seen in patients A, B, H, and I. Within evolution of stable viral haplotypes (clones) is well described for norovirus and influenza in immunocompromised patients *(57,58)* Patients B, H and I were known to be immunocompromised, while Patient A remains under investigation for immune dysfunction. In contrast none of the four adult patients with mixed haplotypes from the independent cohort, were classified as immunocompromised (data not shown). More extensive analysis to understand better the frequency with which within-host variants evolve, why they arise in some patients and not others and whether there are any correlations with underlying clinical condition or outcomes is ongoing.

A major caveat to our findings is that our cohort is paediatric, for which both the clinical picture and outcome of SARS-CoV-2 infections are known to differ from adults. Notwithstanding, similar patterns of clinical and virological response to remdesivir have been described in adults *(3,10,46,59)*. Moreover, both clinical and viral sequence data from the use of repurposed drugs to treat other severe RNA viral infections has shown similarities in adults and children *(12,50,55)*. Larger studies using deep clinical and viral profiling of multiple samples from adult patients treated with remdesivir alone and in combination would provide better insight.

In summary, we show that treatment with remdesivir is capable of reducing SARS-CoV-2 viral and subgenomic RNA *in vivo* and demonstrate that the latter, in particular, needs further investigation as a potential biomarker for monitoring antiviral therapy. Our data suggest that heterogeneous response to remdesivir is likely in many cases to be due to non-viral factors including, potentially, inadequate dosing and duration of treatment. The patterns of SARS-CoV-2 within–host genetic heterogeneity uncovered by deep sequencing may be most parsimoniously explained by viral compartmentalisation within lung-tissue, a factor that is already known to impede drug penetration in patients with other lung infections. This may compound inherently poor remdesivir tissue penetration and rapid clearance of active metabolites in those with normal renal function. We and others have shown that where compartmentalisation occurs in influenza *(29)* and M. *tuberculosis (50)*, combination therapies improve outcomes. Based on our experience of using similar drugs for the treatment of serious RNA viral infections, we propose that a more personalised medicine approach combining *in vitro* and *in vivo* pharmacokinetic measurements, viral RNA and sgRNA profiling together with viral evolutionary modelling is likely to yield important insights into the optimal use of remdesivir both alone and in combination for treatment of SARS-CoV-2. Finally, our data point to the presence of mixed genotypes in a subset of patients, which are most likely explained by within-host evolution rather than co-transmission.

## Materials and Methods

### Study Cohort and approval

We sequenced SARS-CoV-2 samples routinely collected for clinical monitoring from children hospitalized for COVID-19 between early March and mid-May 2020. This study was approved by Great Ormond Street Hospital (Clinical Audit Number #2857) and PHE Research Ethics and Governance Group (REGG) (R&D NR0195).

### Sample Collection and Viral Sequencing

Nasopharyngeal swab samples were collected and tested for SARS-CoV-2. Full length SARS-CoV-2 genome sequences were obtained from all positive samples using SureSelect^XT^ target enrichment and Illumina sequencing. Reads generated were trimmed with trim galore *(60)*. Duplicated reads were removed using Picard *(61)*. For each patient, a unique patient reference was generated by mapping the remaining reads of the first sample to the SARS-CoV-2 reference genome (NC_045512) from GenBank using bwa-mem *(62)*. The mapping quality was checked using Qualimap and the consensus whole genome sequence was generated using QUASR *(63,64)*. Reads from the subsequent samples of the same patient were mapped to this patient reference. Consensus sequences were aligned using MAFFT *(65)*. Only genomes with more than 80 percent genome coverage and a mean read depth of 100 or above were included in downstream analysis. Seqtk was used to subsample the reads for normalisation *(66)*.

### Phylogenetic Analysis

Maximum Likelihood tree of the alignment was constructed using RAxML *(67)*, with the GTR model and 1000 bootstrap replicates. All trees were rooted on the SARS-CoV-2 reference genome NC_045512.

### Analysis and figure generation

Analysis was completed in R 3.6.1 using Rstudio 1.2 *(68,69)*. In general data was processed using dplyr (v0.8.3), figures were generated using ggplot2 (v3.3.1), both part of the tidyverse family of packages (v1.2.1) *(70)*. We employed the fisher.test in R to compare the count-data of mutations for treated and untreated samples across all individuals*(68)*. The Mann-Whitney-Wilcoxon Test was implemented using the wilcox.test in R *(68)*. We used Pearson’s and Spearman’s rank correlation for correlation analysis. This was done using the lm.test and cor.test function in the stats package in R *(68)*.

### Haplotype Reconstruction

Haplotypes were reconstructed using HAplotype Reconstruction Of Longitudinal Deep Sequences (HaROLD) with default settings *(38)*. HaROLD does not statistically support haplotypes where from a single MVA, we therefore constructed by hand the haplotypes for Patient B. In this case the haplotype frequency was taken to be that of the single MVA.

### Quantification of subgenomic RNA

We employed Periscope to detect subgenomic RNA (sgRNA) *(32)*. sgRNA is identified based on the detection of the leader sequence at the 5’end (5’-AACCAACTTTCGATCTCTTGTAGATCTGTTCT-3’) of the sequence.

### Minority Variant Calling

Minority allele variants had to have a frequency of above 2% and with a minimum of 4 supporting reads identified at sites with a read depth of ≥ 5 using VarScan *(71)*.Transient MAVs, which occurred at one timepoint in an individual were discarded from the analysis.

### Glasgow COG-UK Sequencing and Sample Selection for haplotype reconstruction

Minority variants as described above, were called for 318 sequences from Glasgow in the COG-UK from samples collected between 28/02/2020 and 29/04/2020. Following extraction, samples were DNase treated (AM2222) and the libraries were prepared utilising the protocols developed by the ARTIC network (https://artic.network/ncov-2019). Amplicons were generated using either primer sets v1, v2 and v3. The DNA amplicon fragments were cleaned using AMPURE beads (Beckman Coulter) and 40 ng was used to prepare the Illumina sequencing libraries with a DNA KAPA library preparation kit (Roche). Indexing was carried out with NEBNext multiplex oligos (NEB) with 7 cycles of PCR. Libraries were pooled in equimolar amounts and loaded on a MiSeqV2 cartridge (500 cycles). In the case of CR111, the library was sequenced on Illumina’s NextSeq 550 System (Illumina, Part Number SY-415-1002).

To optimise the performance of HaROLD we selected samples with mean read depths of >500 and SNVs occurring at frequencies of >15% with both alleles supported by >100 reads. The number of MVAs was tallied for all samples, and those that reported numbers in the top 2% of all samples were carried forward for analysis by Harold.

### Structural Biology

The structure of the spike protein PDB 6XR8 was visualised using VMD *(72,73)*. Mutations were modelled using Swissmodel *(74,75)*.

## Data Availability

Sequencing data are available upon request.

## Supplementary Materials

### Supplementary Tables

1. Supplementary Table 1: Sequencing Metrics.
2. Supplementary Table 2: Summary of consensus level mutations in patients A, H, and I.
3. Summary of variants detected in all patients.

### Supplementary Figures

1. supplementary figure 1: Patient timeline overview
2. supplementary figure 2: Ct trajectories of 9 patients for all samples collected.
3. supplementary figure 3: Clinical markers for 9 patients.
4. supplementary figure 4: The P812L mutation does not affect the structure of the SARs-CoV-2 spike protein.
5. Supplementary Figure 5: Mutagenic signature for each patient over time.
6. Supplementary Figure 6: Mutational burden over time in each patient.
7. Supplementary Figure 7: Comparison of sgRNA reads per 100,000 mapped reads (sgRPHT) in treated and untreated individuals.
8. Supplementary Figure 8: Polymorphisms trajectories for each patient.
9. Supplementary Figure 9: Comparison of haplotypes from patient A, B, H and I and consensus of C, D, E, G with global sequences.
10. Supplementary Figure 10: Scatter plot of shared polymorphisms (or MVAs) vs pairwise distance for 318 from Glasgow collected between 28/02/2020 and 29/04/2020.
11. Supplementary figure 11: Frequency of identified haplotypes from 4/6 Glasgow samples processed through Harold.

## Acknowledgements

We thank the UK (COG-UK) consortium*. COG-UK is supported by funding from the Medical Research Council (MRC) part of UK Research & Innovation (UKRI), the National Institute of Health Research (NIHR) and Genome Research Limited, operating as the Wellcome Sanger Institute. This work was supported by The John Black Charitable Foundation and the UCLH and GOSH NIHR biomedical research centres. JP is supported by the Rosetrees Foundation and FTB by a Wellcome Trust Collaborative Award to JB. We thank Gilead for supplying Remdesvir on the compassionate access programme and the GOSH bioethics committees together with St Mary’s ID clinicians who contributed to multiplinary discussions relating to compassionate use of RDV. We would like to thank the various medical teams that cared for the patients, with special mention to: Collin Wallis, Rukshana Shroff, Philip Ancliff, Catriona Duncan, Jane Hassel, Guiseppe Barone, Mae Johnson, and Mette Jorgensen. We should also like to acknowledge the contribution of the UCL Pathogen Genomics Unit, UCL Genomics, and the Great Ormond Street Hospital Departments of Infectious Disease, and Microbiology, Virology and Infection Control and Paediatric Intensive Care. This work would not have been possible without the input from patients and their families.

*A full list of consortium names and affiliations can be found at https://www.cogconsortium.uk

