## Supplementary for "Remdesivir induced viral RNA and subgenomic RNA suppression, and evolution of viral variants in SARS-CoV-2 infected patients"

**Supplementary Tables**

**Supplementary Table 1:** Sequencing Metrics. The %OTR and MRD vary across the different samples, with the lowest values associated with low CT values. Samples in orange have low mean read depth all other samples were carried forward for further analysis.

SuppTable1-SequencingMetrics.xlsx

**Supplementary Table 2:** Summary of consensus level mutations in patients A, H, and I relative to their first available sample. No consensus level mutations were found for remaining patients (B, C, D, E, F, and G). Nucleotide replacement relative to entire genome, protein replacement site within gene.

| Patient | Day Post-First Positive | Nucleotide Replacement | Protein Replacement | Gene [product] |
|---|---|---|---|---|
| A | 1, 5, 6, 7 | T10776C | L241P | Orf1ab [nsp5] |
| A | 6, 7, 8 | T9438C | I295T | Orf1ab [nsp4] |
| H | 4,5,6,8,9,10 | T3096C | L126S | Orf1ab [nsp3] |
| H | 4,5,6,8,9,10 | T16308C | synonymous | Orf1ab [nsp13] |
| H | 4,5,6,8,9,10 | G19671A | synonymous | Orf1ab [nsp15] |
| H | 12 | C28253T | synonymous | Orf8 |
| I | 10 | G376T | D37E | Orf1ab [nsp1] |
| I | 10 | C23997T | P812L | S [S2 domain] |
| I | 10 | C28732T | synonymous | N |

**Supplementary Table 3:** Summary of variants detected at 2% reads or above and with at least 4 supporting reads on each strand, relative to each individuals first sample. Haplotype defining sites are highlighted for patients A, B, H, and I.

SuppTable3-Mutations.xlsx

### Supplementary Figures

**Supplementary Figure 1:** Patient timeline overview. First positive are outlined in black, color indicates ward information.

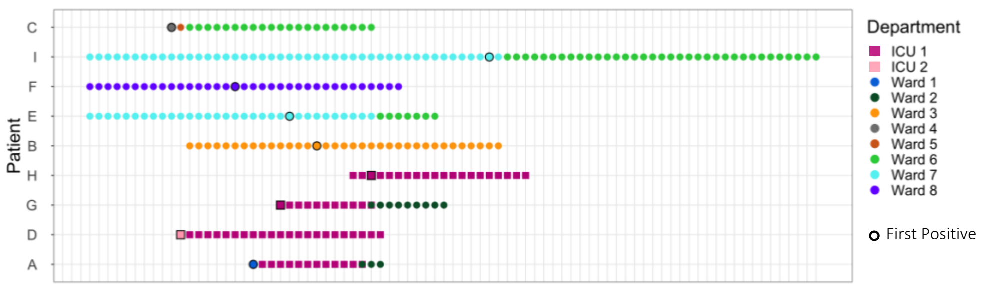

**Supplementary Figure 2:** ct trajectories of 9 patients for all samples collected. One panel per patient, red line indicates remdesivir received, black dot is sample taken, blue circle indicates sample successfully sequenced.

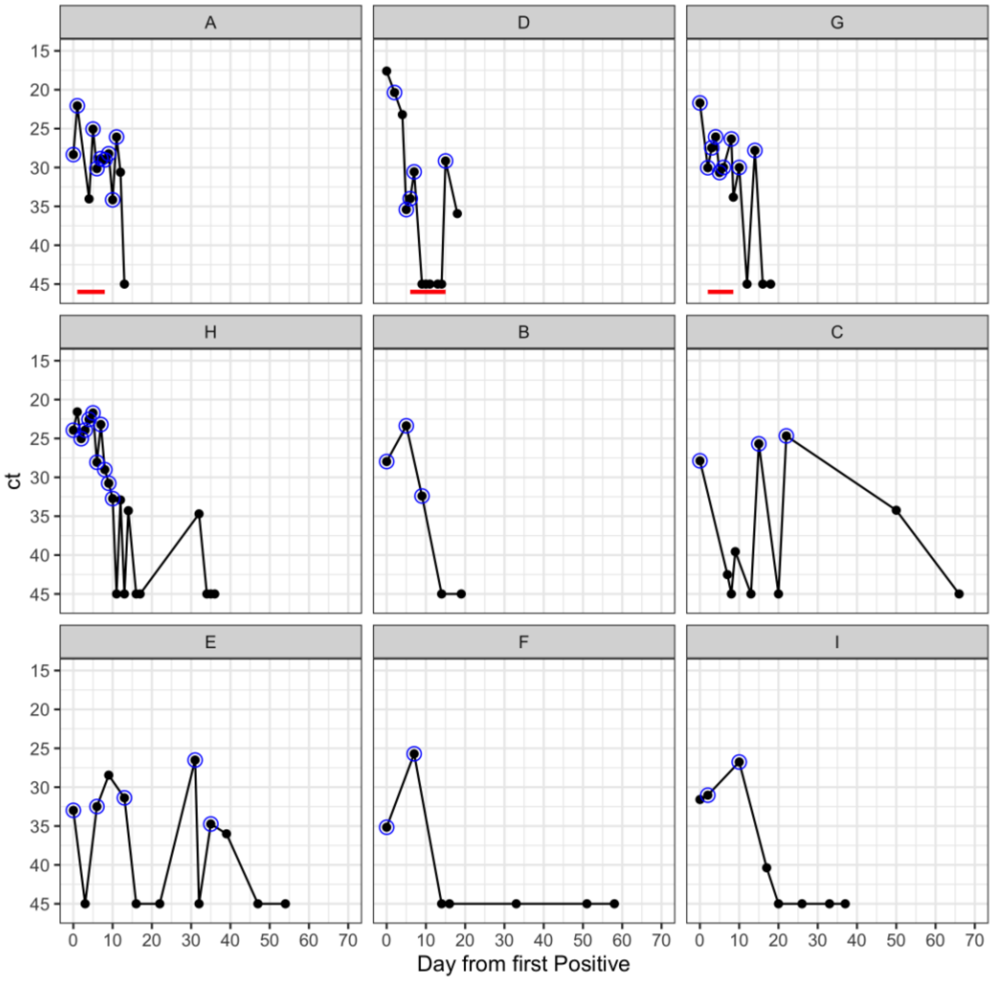

**Supplementary Figure 3:** Clinical markers for 9 patients. Red line indicates administration of remdesivir for patients A, D, and G. Patients A, D, and G all showed resolution of fever after starting remdesivir. In addition, Patients A and G resolved inflammatory markers after starting remdesivir.

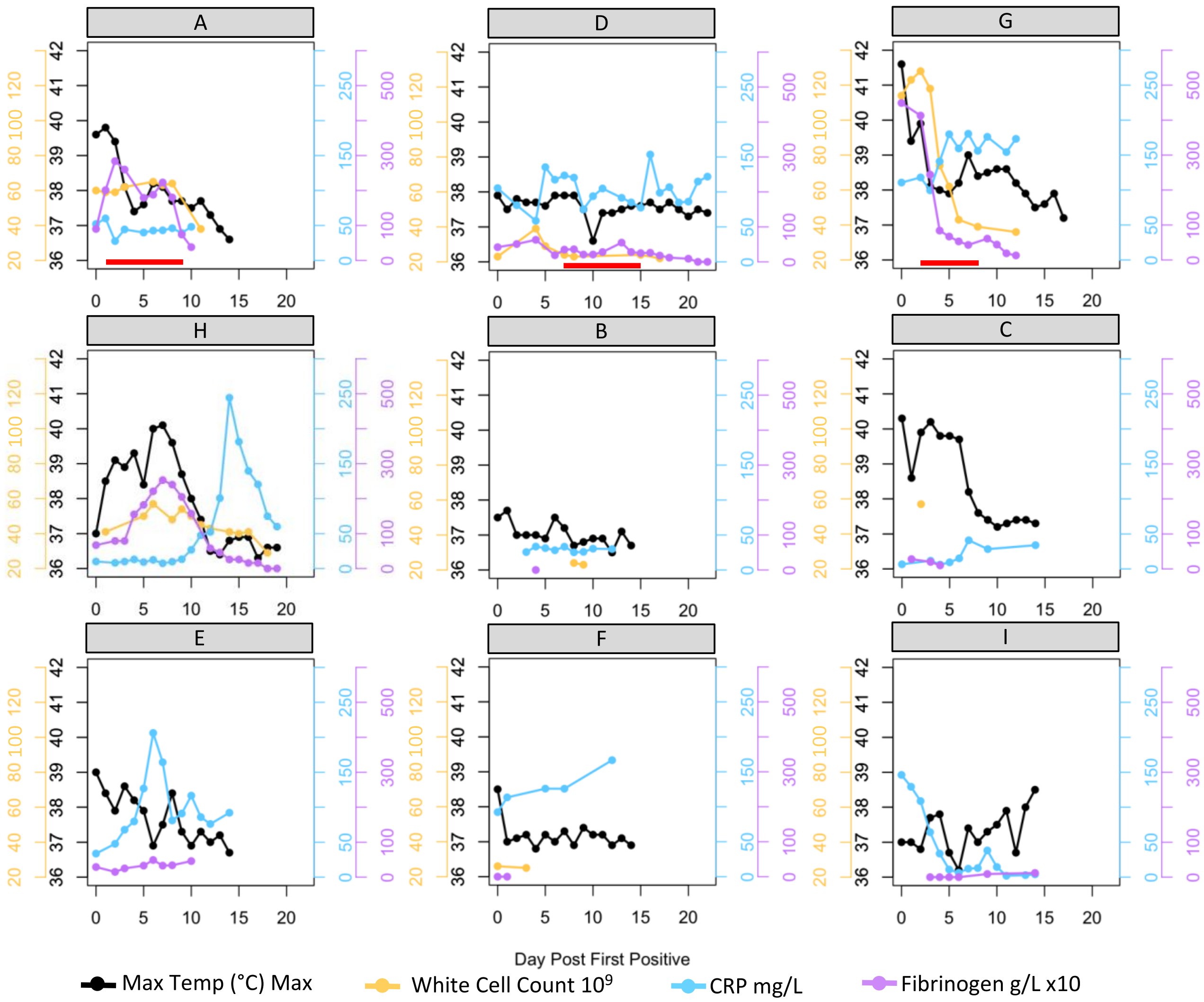

**Supplementary Figure 4:** The P812L mutation does not affect the structure of the SARs-CoV-2 spike protein. A) Spike protein pre-mutation with P812. B) Mutated spike protein with L812 modelled using Swissmodel *(1,2)*. Site 812 is highlighted in orange, the two adjacent residues are highlighted in purple, and the loop is highlighted in blue. The rest of the protein is highlighted in grey.

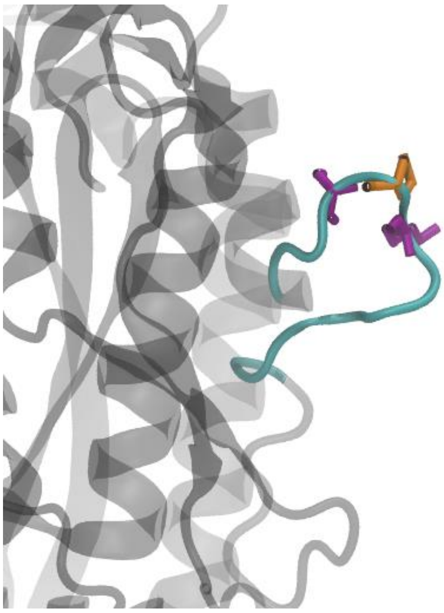

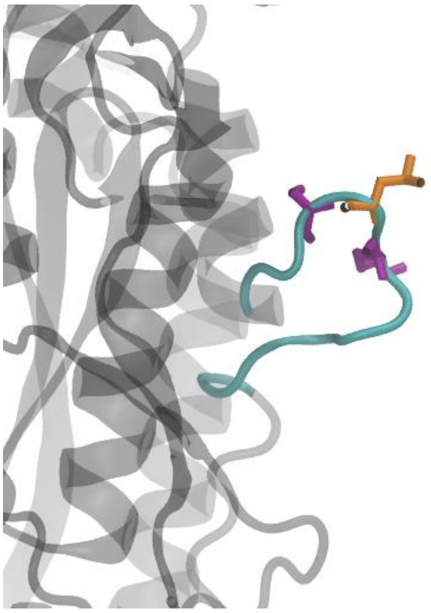

**Supplementary Figure 5:** Mutagenic signature for each patient over time. Stacked bars indicate frequency of transitions and translations.  Red-line indicates administration of remdesivir treatment.

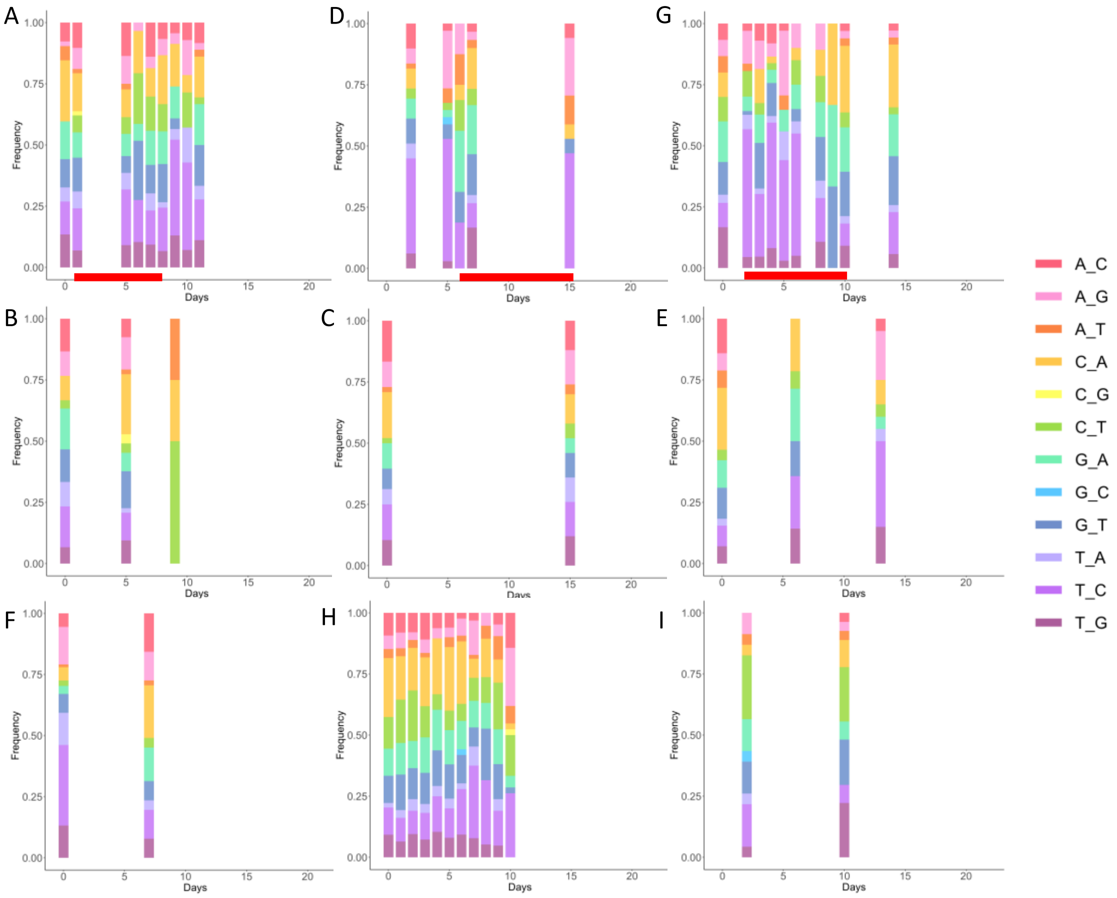

**Supplementary Figure 6:** Mutational burden over time in each patient. X-axis is the count of polymorphisms with frequency higher than 2% from normalized reads as described in the methods. Y-axis is days post-infection. Red line in patients A, D, and G indicate administration of remdesivir. Remdesivir was not found to be associated with a change in mutational burden over time. Mutational burden is relatively stable over time across all patients.

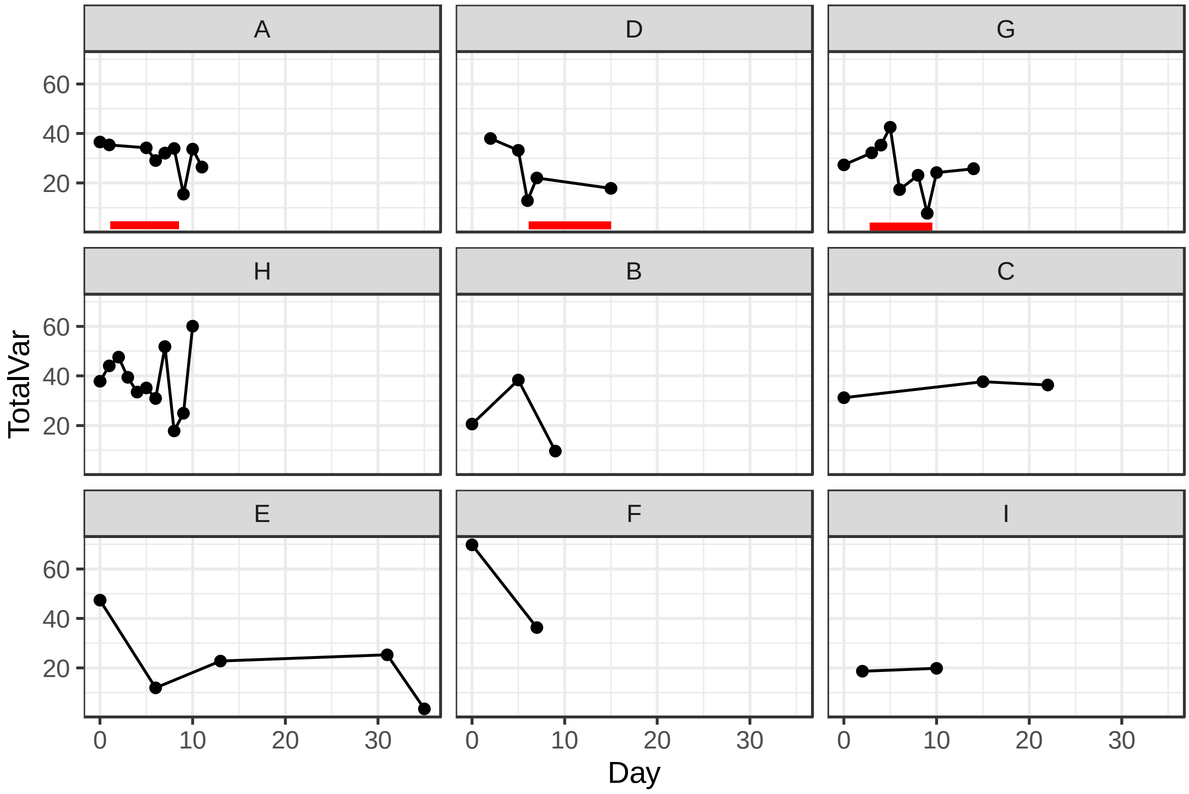

**Supplementary Figure 7:** Comparison of sgRNA reads per 100,000 mapped reads (sgRPHT) in treated and untreated individuals. Each patient is represented by a unique shape and color indicates whether sample point is taken on or off remdesivir. **A)** Scatter-plot of sgRPHT vs ct for all patients. sgRPHT do not correlate with ct values (Spearman-rank-correlation rho=-0.206, p =0.156). **B)** box-plot comparing sgRPHT in treated and untreated samples across entire dataset. Patient D is excluded as we do not have measurement of sgRPHT on remdesivir. There is a tendency towards statistical significance change in sgRPHT associated with remdesivir treatement when comparing all samples on and off remdesivir independent to time of infection (Mann-Whitney-Wilcoxon test, p = 0.059).

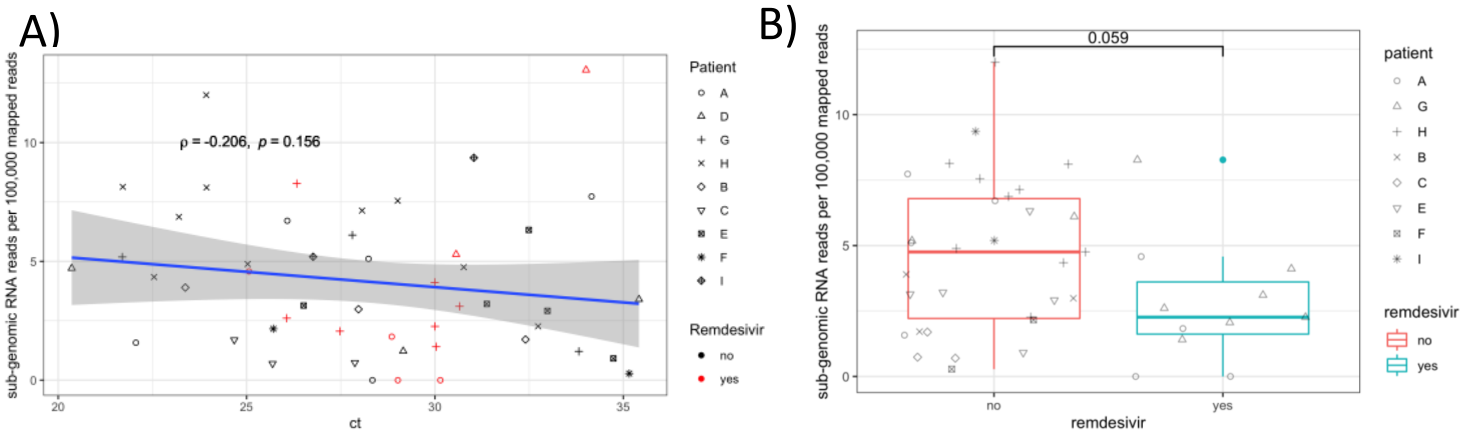

**Supplementary Figure 8:** Polymorphisms trajectories for each patient. *Top panel*: ct values red line indicates remdesivir received, black dot is sample taken, blue circle indicates sample successfully sequenced. *Bottom panel:* Variant reads frequencies color coded by site, in grey scale are transient polymorphisms.

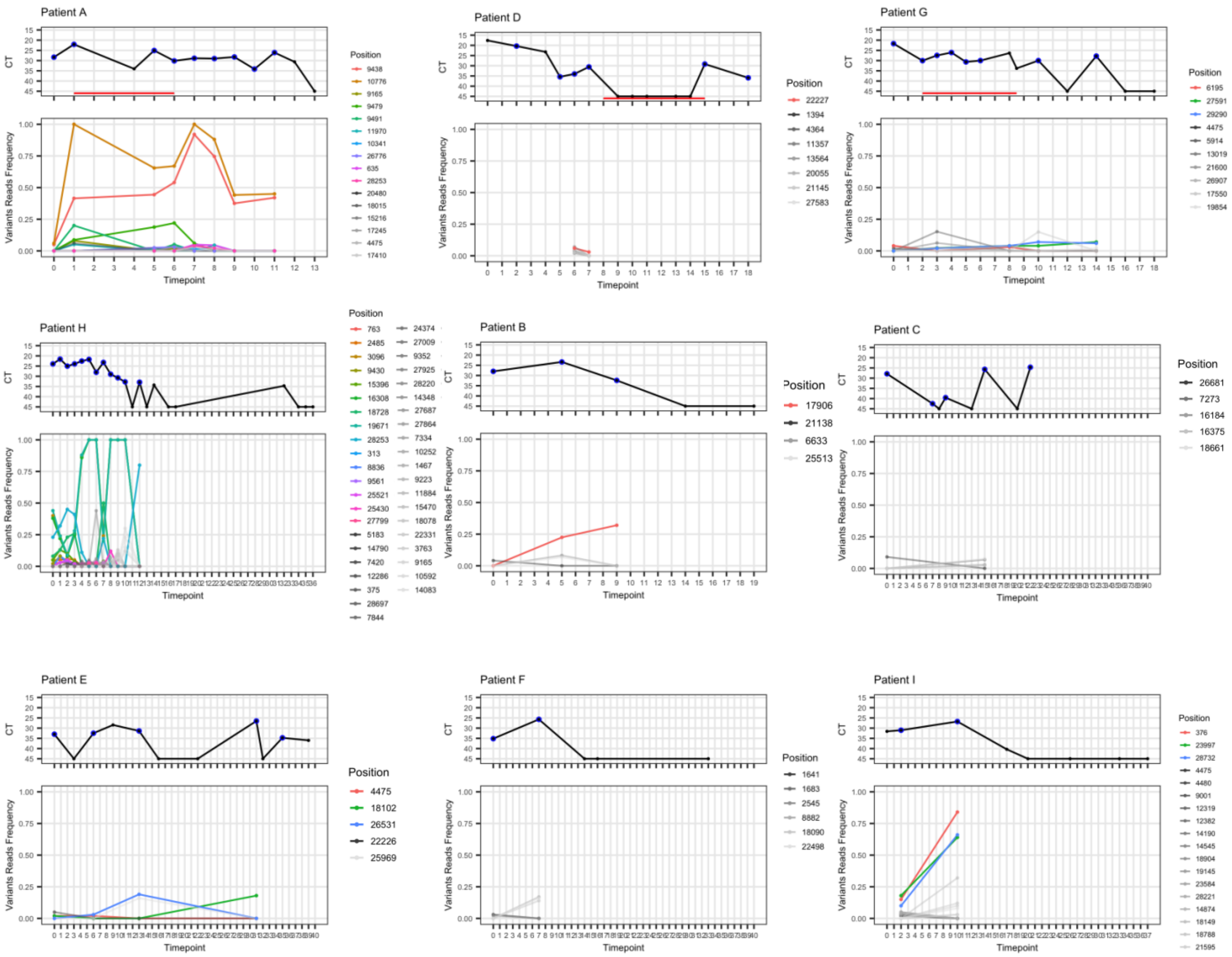

**Supplementary Figure 9:** Comparison of haplotypes from patient A , B, H, and I and consensus of C, D, E, G with global sequences. Local tree representing closest samples from global database for haplotpyes in patients A, H and I. Trees are grouped by corresponding lineages for Patients C, D, E, F, and G.

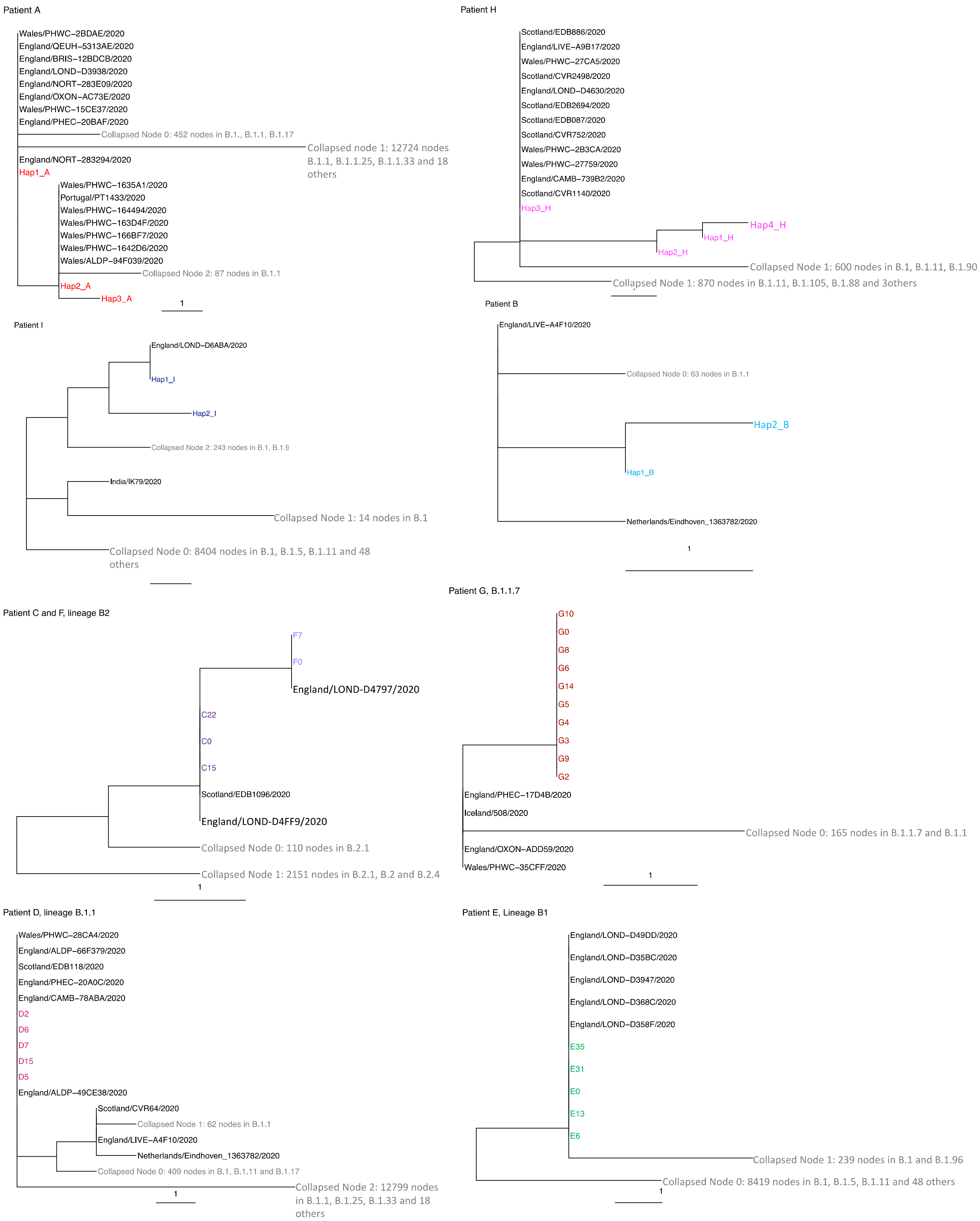

**Supplementary Figure 10:** Scatter plot of shared polymorphisms (or MVAs) vs pairwise distance for 318 from Glasgow collected between 28/02/2020 and 29/04/2020. Size of dot indicates number of pairs of samples compared.  No correlation is found between shared variants and pairwise distance (Pearson's correlation $r^2$ = 0.00242).

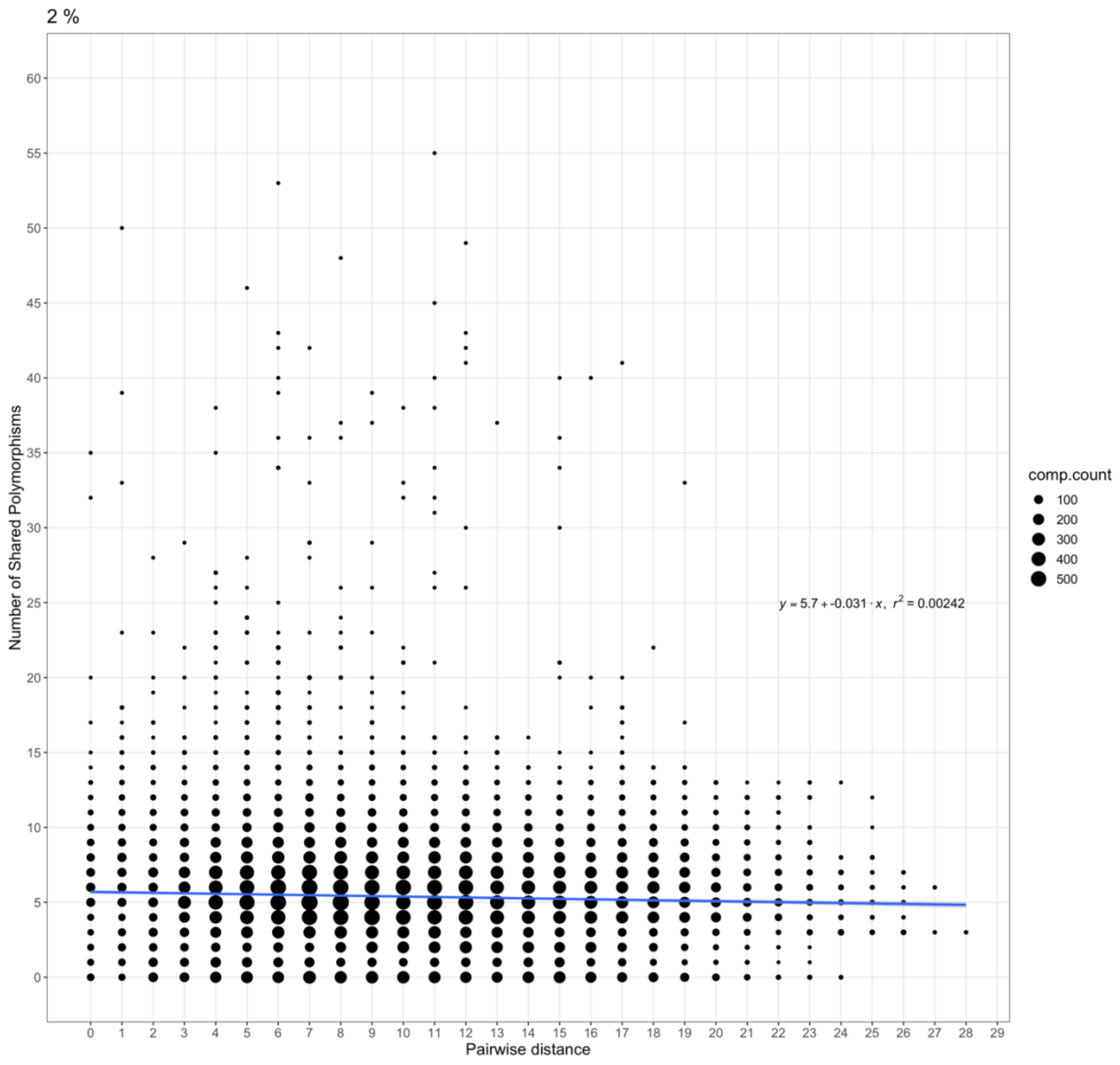

**Supplementary figure 11**: Frequency of identified haplotypes from 4/6 Glasgow samples processed through Harold. HaROLD identifies between 2-3 distinct haplotypes for each sample.

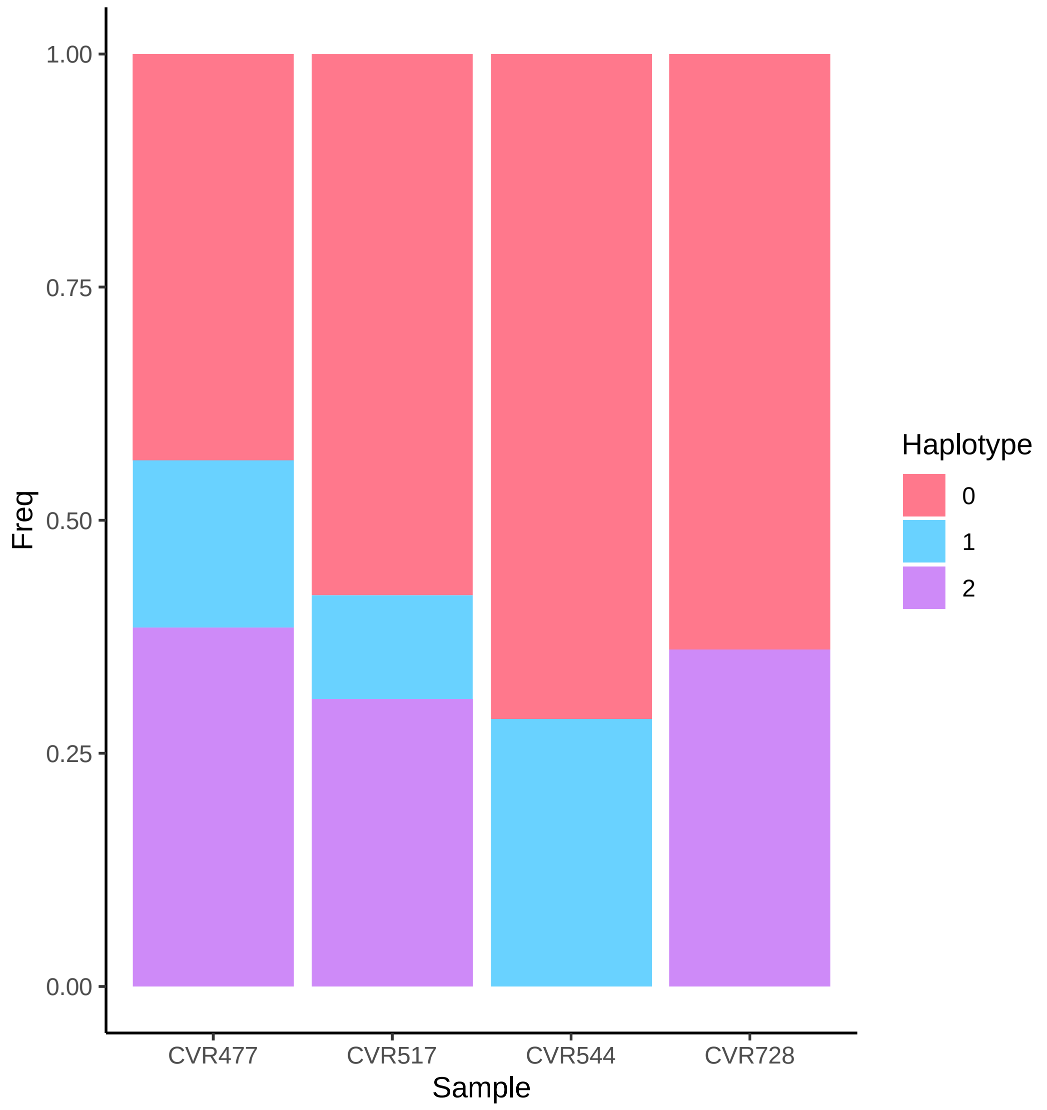
